# Omicron BA.1 and BA.2 Neutralizing Activity Following Pre-Exposure Prophylaxis with Tixagevimab plus Cilgavimab in Vaccinated Solid Organ Transplant Recipients

**DOI:** 10.1101/2022.05.24.22275467

**Authors:** Andrew H. Karaba, Jake D. Kim, Teresa P-Y Chiang, Jennifer L. Alejo, Aura T. Abedon, Jonathan Mitchell, Amy Chang, Yolanda Eby, Trevor Scott Johnston, Tihitina Aytenfisu, Casey Hussey, Alexa Jefferis, Nicole Fortune, Rivka Abedon, Letitia Thomas, Daniel S. Warren, Ioannis Sitaras, Andrew Pekosz, Robin K. Avery, Allan B. Massie, William A. Clarke, Aaron A.R. Tobian, Dorry L. Segev, William A. Werbel

## Abstract

Neutralizing antibody responses are attenuated in many solid organ transplant recipients (SOTRs) despite SARS-CoV-2 vaccination. Pre-exposure prophylaxis (PrEP) with the monoclonal antibody combination Tixagevimab and Cilgavimab (T+C) might augment immunoprotection, yet activity against Omicron sublineages in vaccinated SOTRs is unknown. Vaccinated SOTRs who received 300+300mg T+C (either single dose or two 150+150mg doses) within a prospective observational cohort submitted pre- and post-injection samples between 1/10/2022-4/4/2022. Binding antibody (anti-receptor binding domain [RBD], Roche) and surrogate neutralization (%ACE2 inhibition; ≥20% connoting neutralizing inhibition, Meso Scale Discovery) were measured against variants including Omicron sublineages BA.1 and BA.2. Data were analyzed using the Wilcoxon matched-pairs signed-rank test and McNemar’s test. Among 61 participants, median (IQR) anti-RBD increased from 424 (IQR <0.8-2322.5) to 3394.5 (IQR 1403.9-7002.5) U/ml post T+C (p<0.001). The proportion demonstrating vaccine strain neutralizing inhibition increased from 46% to 100% post-T+C (p<0.001). BA.1 neutralization was low and did not increase (8% to 16% of participants post-T+C, p=0.06). In contrast, BA.2 neutralization increased from 7% to 72% of participants post-T+C (p<0.001). T+C increased anti-RBD levels, yet BA.1 neutralizing activity was minimal. Encouragingly, BA.2 neutralization was augmented and in the current variant climate T+C PrEP may serve as a useful complement to vaccination in high-risk SOTRs.

## 1. Introduction

Many solid organ transplant recipients (SOTRs) exhibit poor binding antibody response and plasma neutralizing capacity against SARS-CoV-2 variants of concern (VOC) despite repeated vaccine doses^1-3^. This, in part, underlies higher rates of clinically significant infection after vaccination (“breakthrough”) in SOTRs^4,5^, and recommendations for additional vaccine doses. The injectable monoclonal antibody combination Tixagevimab and Cilgavimab (T+C) was recently authorized by the US Food and Drug Administration (FDA) as pre-exposure prophylaxis (PrEP) versus SARS-CoV-2, a complementary strategy to reduce COVID-19 in immunocompromised persons^6^. Supporting data, however, were based on trials of unvaccinated, immunocompetent persons and preceded the rise of the Omicron variant and its sublineages, which exhibit significant immune evasion^7,8^. Indeed, *in vitro* data indicated a >100-fold decrease in neutralization of BA.1 and supported doubling of the recommended dose of T+C from 150+150mg (150mg Tixagevimab and 150mg Cilgavimab) to 300+300mg (300mg Tixagevimab and 300mg Cilgavimab) ^9^. Due to uncertainty regarding the protection afforded by T+C, we analyzed anti-spike receptor binding domain (RBD) antibody responses and plasma neutralizing capacity against VOC including Omicron sublineages as well as tolerability in a real-world observational cohort of vaccinated SOTRs.

## 2. Methods

### 2.1 Cohort

SOTRs were enrolled in a national, prospective observational study of SARS-CoV-2 vaccine response (Johns Hopkins IRB00248540) as previously described ^10,11^. All participants were contacted in January 2022 to report prior or planned receipt of T+C (150+150mg dose), with a second communication in March 2022 to capture receipt of 300+300mg dosing (either as two 150+150mg doses or one as 300+300mg dose) following revised FDA recommendations. Participants were enrolled and consented electronically. All participants receiving a total of 300+300mg dosing (i.e., full dose) between January 10 and April 4, 2022 were included in this analysis (n=61). The study team neither administered T+C nor encouraged its receipt, and the doses were independently administered in the community. Participants were stratified by history of recent SARS-CoV-2 antigen exposure, defined as vaccination between 30 days prior to first T+C injection and first post-T/C sample collection date, or SARS-CoV-2 infection between 90 days prior to first T+C injection and first post-T/C sample collection date, as this may have confounded changes in immunogenicity measurements.

### 2.2 Sample collection and processing

Participants provided whole blood samples via at-home phlebotomy service ≤2 weeks before and two weeks following each T+C dose. Blood was collected in 8.5mL acid citrate dextrose tubes and shipped overnight to the study team (Johns Hopkins University). Plasma was separated via centrifugation and stored at –80°C.

### 2.3 Binding antibody measurements

Plasma samples were tested on two anti-spike assays (one clinical, one research) given a priori uncertainty regarding the capture of the monoclonal antibody product. This included the clinical Roche Elecsys anti-SARS-CoV-2-S anti-receptor binding domain (RBD) pan immunoglobulin assay (units/mL, approximately 1:1 with World Health Organization binding antibody units [BAU]). Samples with anti-RBD >250 U/mL were successively diluted until signal fell within quantification range. Additionally, the Meso Scale Diagnostics (MSD, Rockville, MD) research assay was used to measure anti-RBD and anti-nucleocapsid (anti-N) binding antibody via the V-PLEX COVID-19 Respiratory Panel 3 Kit at 1:5000 dilution per manufacturer’s protocol. Plates were read on a MESO QuickPlex SQ 120 and arbitrary units (AU) were calculated using the MSD Discovery Workbench software according to the manufacturer’s protocol. Conversion to WHO binding antibody units (BAU) was done by multiplying by the manufacturer’s recommended conversion factor. The upper limit of quantification (ULOQ) for MSD anti-RBD assay is 4500 BAU. The imputed value from the manufacturer’s software was used if a signal was above ULOQ.

### 2.4 Surrogate neutralization (percent ACE2 inhibition)

The MSD chemiluminescent assay was used to measure the inhibition of ACE2 receptor binding to the spike protein (%ACE2 inhibition) as previously described^3^. Samples were assayed on MSD SARS-CoV-2 panel 25 at a dilution of 1:100 and tested against the ancestral strain (vaccine), B.1.1.7 (Alpha), B.1.351 (Beta), B.1.617.2 (Delta), BA.1 (Omicron), and BA.2 (Omicron). Based on prior work in SOTRs utilizing authentic live virus assays, ≥20% ACE2 inhibition was defined as “neutralizing inhibition” given the consistent association with live virus neutralizing capacity in this specific immunosuppressed population^3^.

### 2.5 Statistical analysis

Wilcoxon matched-pairs signed-rank test was used to compare the anti-RBD level pre- and post-T+C. McNemar’s test was used to compare the frequency of achieving neutralizing inhibition (≥20%) pre- and post-T+C. Analyses were repeated for the pre-specified subgroup without recent SARS-CoV-2 antigen exposure. For SOTRs receiving two 150+150mg doses of T+C, the longitudinal trajectory of anti-RBD and %ACE2 inhibition were measured after each dose to quantify the effect of a second 150+150mg dose. The correlation between the log10-transformed clinical and research assays, as well as the clinical assay and percent ACE2 inhibition were examined using Spearman’s rank correlation coefficients^12^. Scatterplots of % ACE2 inhibition versus anti-RBD were visualized to assess potential thresholds associated with neutralizing inhibition, by variant. Analyses were conducted on Stata/SE 17.0 (College Station, TX) and Microsoft Excel (2019).

### 2.6 Safety assessments

Participants received electronic surveys 7 days following each T+C injection to solicit adverse events, specifically querying for cardiac events and hypersensitivity reactions. The survey also queried for local (pain, redness, swelling) and systemic (fever, fatigue, headache, diarrhea, myalgia, chills, and vomiting) symptoms. Symptoms were graded as mild (“does not interfere with activity”); moderate (“some interference with activity”); or severe (“prevents daily activity”). Breakthrough SARS-CoV-2 infections were defined as either self-reported new diagnosis of COVID-19 or anti-N seroconversion. Breakthroughs were ascertained by surveys at 7 days following each T+C injection, as well as via unsolicited participant self-report. As part of the larger COVID-19 observational study of SARS-CoV-2 vaccine response, participants were highly encouraged to report COVID-19 diagnosis or adverse events via multiple communications throughout the span of the study.

## 3. Results

### 3.1: Demographics and vaccination history

Of 61 included participants, 21 received a single 300+300mg T+C dose and 40 received two 150+150mg doses. Median (interquartile range [IQR]) age was 62.5 (57.7-68.5) years, 36 (59%) participants were female, and 7 (11%) were non-white. The majority, 32 (52%), were kidney transplant recipients and 16 (26%) were thoracic transplant recipients (8 [13%] heart and 8 [13%] lung). The median time from transplant to T+C administration was 4.9 (2.2-11.1) years. For participants receiving a single 300+300mg T+C, the median follow-up time was 13 (7, 15) days post-T/C. For participants receiving two 150+150mg T+C, the median follow-up time was 45.5 (29, 61.5) days and 13.5 (7.5, 25) days post first and second 150+150mg T+C injection, respectively. The majority, 32 (52%), reported taking triple immunosuppression, defined as calcineurin inhibitor, antimetabolite, and corticosteroids. All participants had completed at least a three-dose primary series of SARS-CoV-2 vaccination pre-T+C, including 38 who received four doses and 5 who received five doses. All participants received either an mRNA vaccine (BNT162b2 or mRNA-1273) or Ad.26.COV2.S as a third dose. 54/61 (88%) participants received mRNA vaccine for all three doses of primary series; 36/38 (95%) and 5/5 (100%) participants received mRNA vaccine as fourth and fifth dose, respectively. Of the 22 participants stratified by pre-defined SARS-CoV-2 antigen exposure, 16 received SARS-CoV-2 vaccine <30 days pre-T+C, 3 received SARS-CoV-2 vaccine after T+C injection, and 3 reported SARS-CoV-2 infection <90 days pre-T+C (**Table 1**).

**Table 1.** Clinical and demographic characteristics of SOTRs receiving Tixagevimab plus Cilgavimab

|  | Overall (N=61) | Recent<br>Vaccination or<br>COVID-19 (N=22) | No Recent<br>Vaccination or<br>COVID-19 (N=39) |
| --- | --- | --- | --- |
| Age, years (%) |  |  |  |
| 20-39 | 3 (5) | 2 (9) | 1 (3) |
| 40-59 | 20 (33) | 7 (32) | 13 (33) |
| 60-79 | 38 (62) | 13 (59) | 25 (64) |
| Female sex (%) | 36 (59) | 9 (41) | 27 (69) |
| Nonwhite race (%) | 7 (11) | 2 (9) | 5 (13) |
| Years since transplantation (IQR) | 4.9 (2.2-11.1) | 5.8 (2.3-11.7) | 4.3 (2.1-11.1) |
| Graft transplanted (%) |  |  |  |
| Kidney | 32 (52) | 9 (41) | 23 (59) |
| Liver | 5 (8) | 0 (0) | 5 (13) |
| Heart | 8 (13) | 5 (23) | 3 (8) |
| Lung | 8 (13) | 6 (27) | 2 (5) |
| Multi-organ | 8 (13)<sup>1</sup> | 2 (9)<sup>2</sup> | 6 (15)<sup>3</sup> |
| Anti-rejection medications (%)<sup>4</sup> |  |  |  |
| Triple immunosuppression | 32 (52) | 11 (50) | 21 (54) |
| CNI | 54 (88) | 20 (91) | 34 (87) |
| Steroids | 41 (67) | 14 (64) | 27 (69) |
| Mycophenolate | 52 (85) | 18 (82) | 34 (87) |
| mTOR | 8 (13) | 5 (23) | 3 (8) |
| Tacrolimus | 49 (80) | 17 (77) | 32 (82) |
| Belatacept | 2 (3) | 0 (0) | 2 (5) |
| Days since last vaccination (IQR) | 82 (26-161) | 20 (15-27) | 116 (44-161) |
| Number of Vaccine Doses pre-T+C (%) |  |  |  |
| Three | 23 (38) | 4 (18) | 19 (49) |
| Four | 33 (54) | 17 (77) | 16 (41) |
| Five | 5 (8) | 1 (5) | 4 (10) |
| Received mRNA vaccine (%)<sup>5</sup> |  |  |  |
| Primary series<sup>5</sup> | 54/61 (88) | 22/22 (100) | 32/39 (82) |
| Fourth Dose | 40/42 (95) | 20/20 (100) | 20/22 (91) |
| Fifth Dose | 5/5 (100) | 2/2 (100) | 3/3 (100) |
| Prior COVID-19 (%)<sup>6</sup> | 3 (5) | 3 (14) | 0 (0) |
| Recent COVID-19 vaccination (%)<sup>7</sup> | 20 (33) | 20 (91) | 0 (0) |

<sup>1</sup> 2 kidney-liver, 4 kidney-pancreas, 1 kidney-heart, 1 lung-intestine
<sup>2</sup> 1 kidney-heart, 1 lung-intestine
<sup>3</sup> 2 kidney-liver, 4 kidney-pancreas
<sup>4</sup> anti-rejection medications are not mutually exclusive
<sup>5</sup> Received mRNA all three doses
<sup>6</sup> within 90 days pre-T+C
<sup>7</sup> one participant received vaccination post-T+C
Abbreviations: CN1 calcineurin inhibitor; mTOR mammalian target of rapamycin inhibitor

### 3.2: Change in binding antibody

Following a full dose T+C, median (IQR) anti-RBD titer increased from 424 (<0.8, 2323) U/ml to 3500 (1433, 7115) U/ml on the clinical assay (p<0.001) (**Figure 1**); 16 (26%) participants had no detectable antibody on the clinical assay prior to T+C injection, all of whom seroconverted. The change in titer was similar for those receiving a single 300+300mg dose versus two 150+150mg doses (**Figure S1**). Anti-RBD titer on the research assay similarly increased, from median (IQR) 336.7 (8.1, 2858) to 8185 (6253, 10658) BAU (p<0.001) (**Table S1A**). There was similar change in anti-RBD titer among participants without recent antigen exposure (n=39), from median (IQR) anti-RBD 393 (<0.8, 3453) U/mL to 3638 (1363, 7115) U/mL on the clinical assay (p<0.001), and from 352.7 (IQR 10.2, 3422) BAU to 8029 (4894, 10658) BAU on the research assay (p<0.001) (**Table S1B**). There was a strong positive correlation between the clinical and research assay titers for all values below the upper limit of quantification of 4500 BAU (Spearman ρ=0.84, 76 samples, n=51) (**Figure S2**).

**Figure 1.**
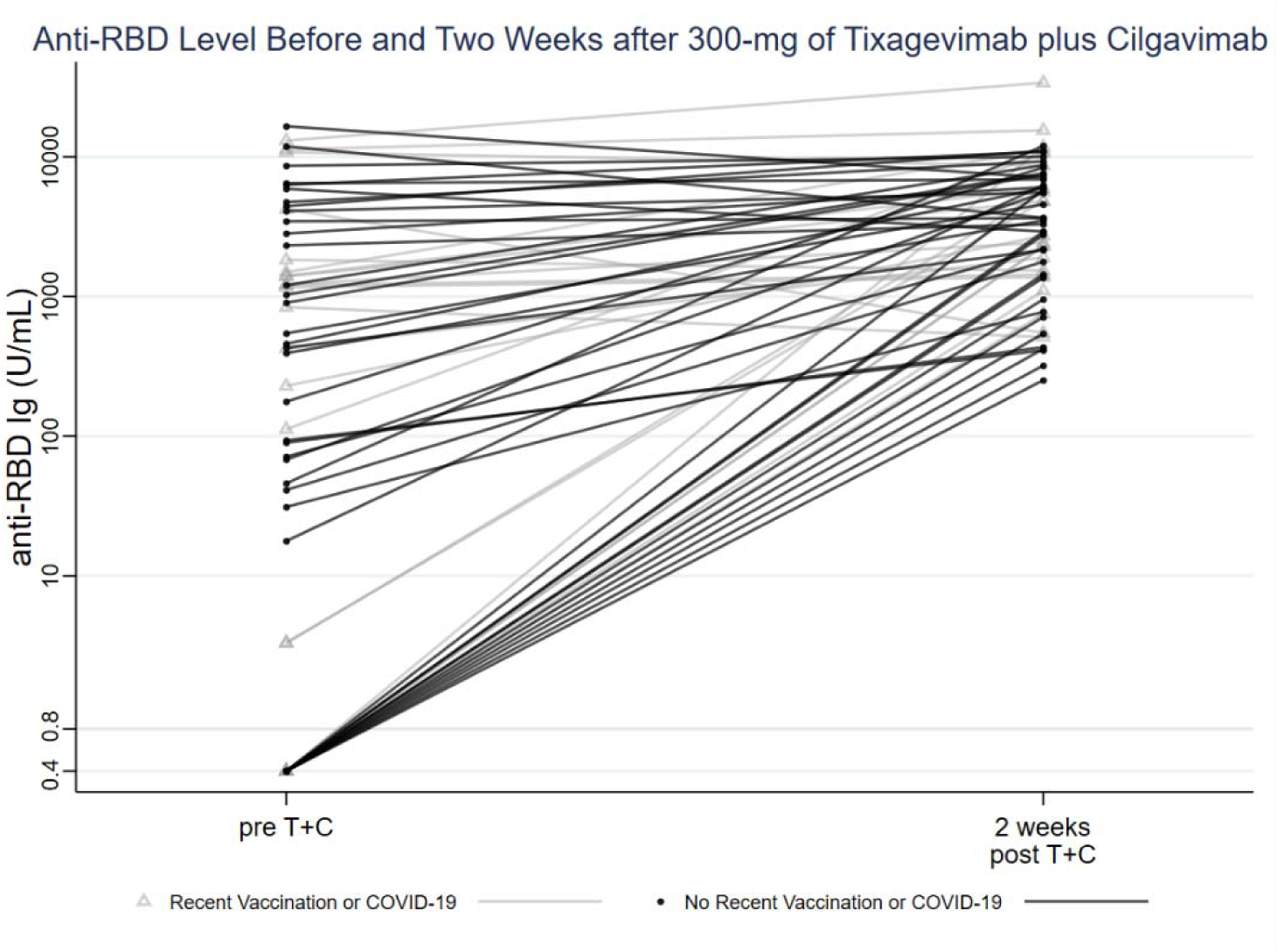
Anti-RBD level before and two weeks after 300+300mg of Tixagevimab plus Cilgavimab

### 3.3: Change in surrogate neutralization

The proportion of participants who exhibited neutralizing inhibition (≥20%) against the vaccine strain increased from 46% (28/61) pre-T+C to 100% (61/61) post full-dose T+C (Exact McNemar p<0.001). All participants exhibited neutralizing inhibition after T+C against the Alpha variant (44% to 100%, p<0.001), Beta variant (26% to 100%, p<0.001) and Delta Variant (39% to 100%, p<0.001) (**Tables S2A and S3A, Figure S3**).

In contrast, the proportion of participants who exhibited neutralizing inhibition against the Omicron BA.1 sublineage did not significantly increase: 8% (5/61) pre-T+C versus 16% (10/61) post full-dose T+C (p=0.06). For the Omicron BA.2 sublineage, however, the proportion who exhibited neutralizing inhibition increased from 7% (4/61) to 72% (44/61) post-T+C (p<0.001). Among the 39 participants without recent antigen exposure, changes in surrogate neutralization post-T+C were similar against tested variants (**Table S2B and S3B, Figure 2**).

**Figure 2.**
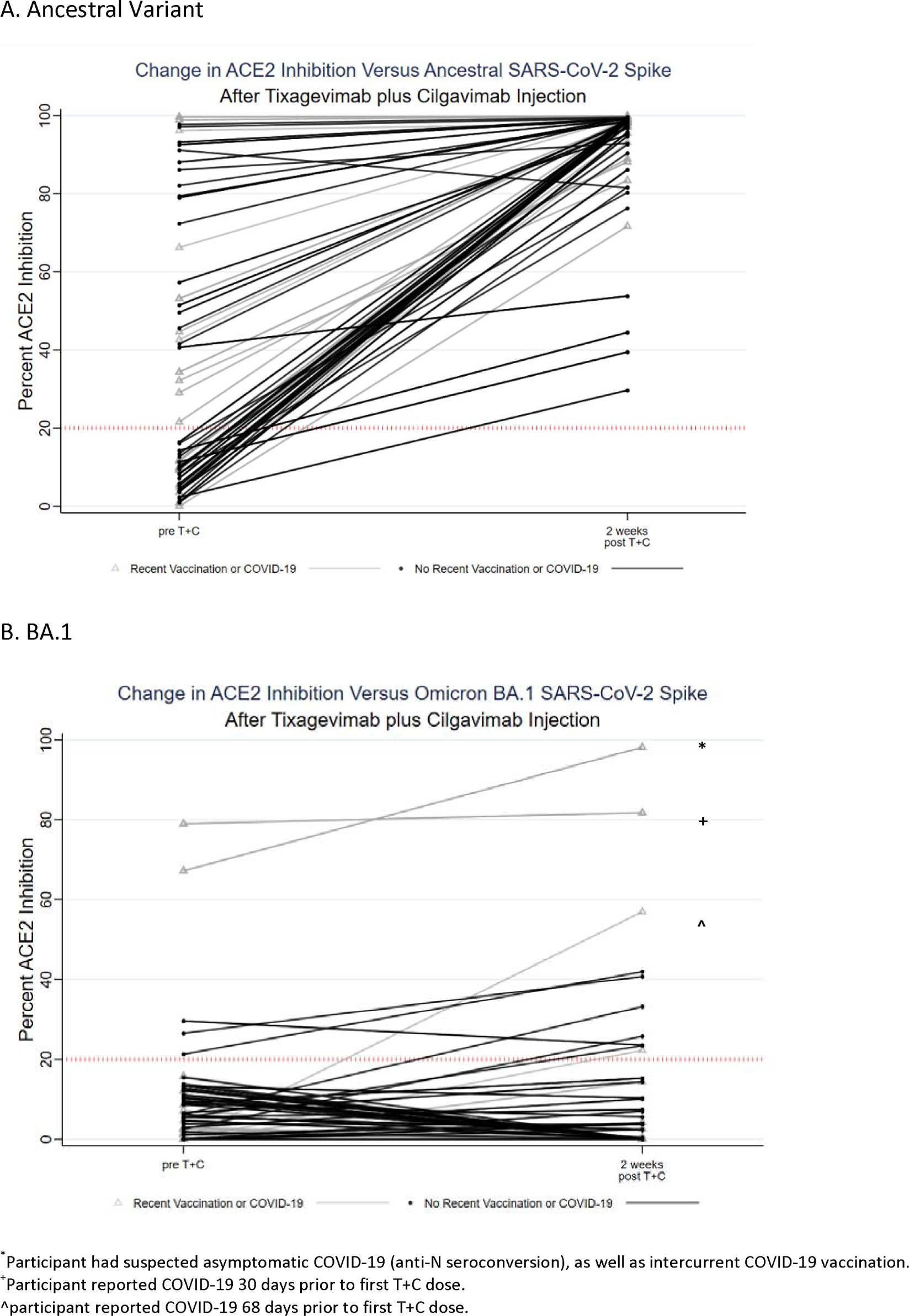
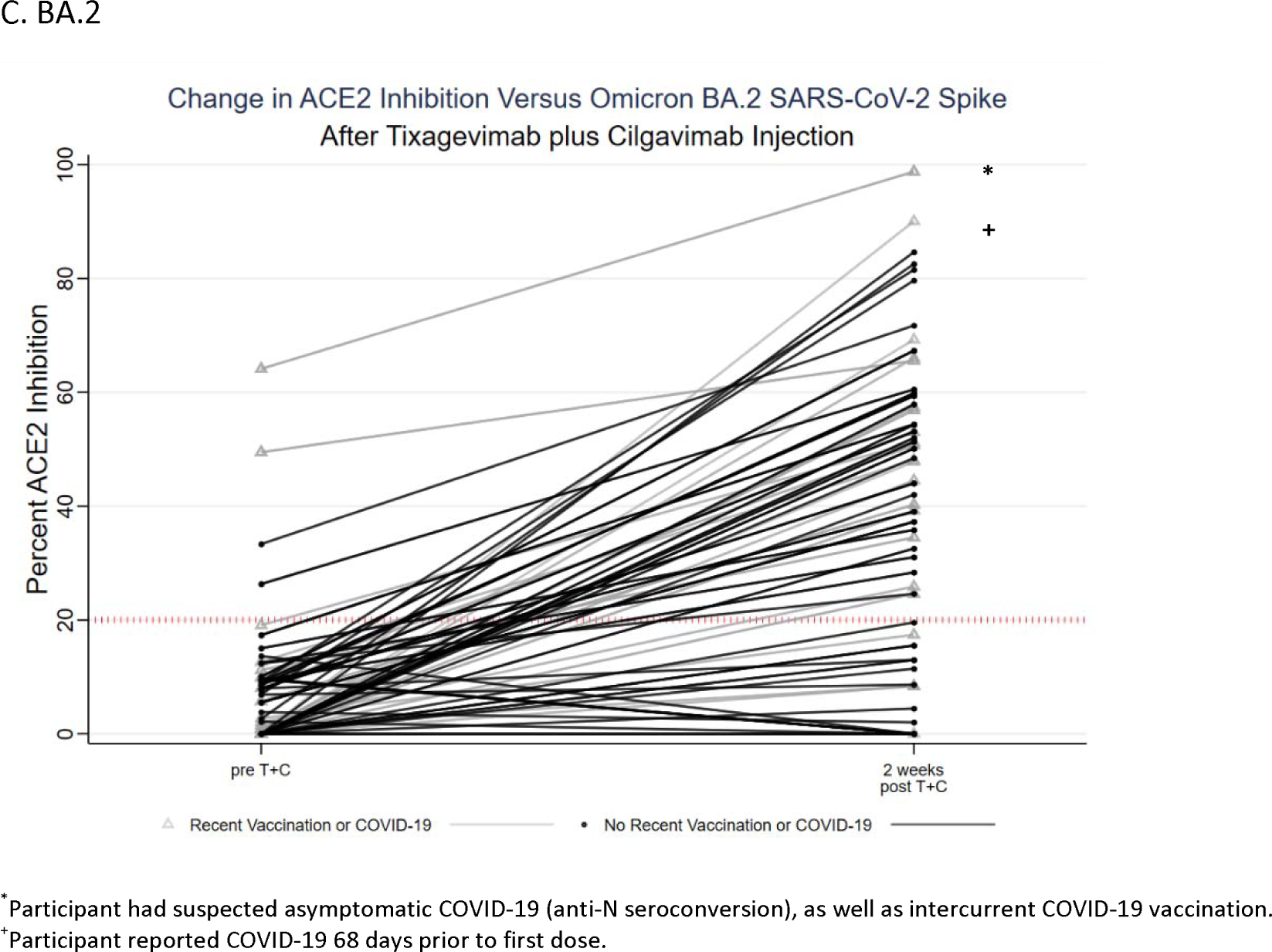
Change in ACE2 inhibition versus SARS-CoV-2 variants after 300+300mg of Tixagevimab plus Cilgavimab injection

### 3.4 Association between binding antibody and surrogate neutralization

There was a moderate positive correlation between anti-RBD titer and %ACE2 inhibition against the vaccine strain, alpha, beta, and delta variants (vaccine: ρ=0.65, alpha: ρ=0.62, beta: ρ=0.61, delta ρ=0.62). There was an attenuated correlation between anti-RBD and %ACE2 inhibition for Omicron sublineages, though most SOTRs who achieved neutralizing inhibition had high levels of anti-RBD after T+C: median (IQR) 7888 (2433, 10865) (n=10) for BA.1 and 4696 (2027, 8688) for BA.2 (n=44) (**Figure S4, Table S4**).

### 3.5 Impact of a second 150+150mg T+C dose on VOC neutralization

Among 40 participants who received two 150+150mg doses, the proportion reaching neutralizing inhibition against the vaccine strain increased from 43% (17/40) pre-T+C, to 97% (32/33) post-T+C dose 1, and 100% (40/40) post-T+C dose 2. The proportion reaching neutralizing inhibition against Omicron BA.1 did not increase: 10% (4/40) pre-T+C, to 6% (2/33) post-T+C dose 1, and 10% (4/40) post-T+C dose 2. However, the proportion reaching neutralizing inhibition against Omicron BA.2 increased from 8% (3/40) pre-T+C, to 55% (18/33) post-T+C dose 1, and 80% (32/40) post-T+C dose 2 (**Figure S5**).

### 3.6 Safety and SARS-CoV-2 infections

Zero participants reported an adverse cardiac event, hypersensitivity reaction, or acute organ rejection within 7 days of any T+C injection. Reported reactions were mild or moderate and were more frequent after 300+300mg dosing versus 150+150mg dosing (**Figure S6**). There were no reported symptomatic breakthrough SARS-CoV-2 infections, though one participant showed anti-N seroconversion after a first 150+150mg dose of T+C, consistent with asymptomatic infection. The median (IQR) of follow-up after T+C was 13 (7, 19).

## 4. Discussion

This real-world observational study of vaccinated SOTRs receiving T+C indicates that monoclonal antibody-based PrEP increases binding antibody responses and plasma neutralizing capacity against earlier VOCs, yet confirms variable effect against Omicron sublineages. Specifically, there was no observed improvement in BA.1 neutralization, with nearly 85% of recipients lacking neutralizing inhibition two weeks post-injection. In contrast, neutralizing activity against BA.2, the current dominant sublineage in the United States and Europe ^13^, was augmented; 72% of T+C recipients reached neutralizing inhibition. In keeping with updated FDA recommendations, the impact of the increased 300+300mg dose was pronounced and may be necessary to achieve BA.2 neutralization for many SOTRs. Reassuringly, T+C was well-tolerated and there were no early signals for serious adverse events including cardiac complications as described in the landmark PROVENT trial, though a future study of a longer timeframe would be necessary to determine longitudinal adverse events in SOTRs^7^.

Our data suggest that T+C is a reasonable adjunct to vaccination in SOTRs, yet may not provide universal protection against SARS-CoV-2 infection amid an evolving variant climate. Indeed, preliminary data indicates that breakthrough COVID-19 during the Omicron era has occurred in SOTRs despite full vaccination and T+C injection, particularly with BA.1 sublineages^14^. Furthermore, long-term breakthrough post-TC against BA.2 needs to be investigated given the current dominance of BA.2 sublineage.

It is notable that common binding antibody assays are capable of detecting changes in anti-RBD levels following T+C injection and provide similar results. Consistent with prior work^15^, high levels of binding antibody were associated with Omicron neutralizing inhibition, which supports the role of antibody testing as one component of clinical decision-making regarding risk stratification and the timing of additional active or passive immunization (i.e., booster vaccinations and/or repeat T+C dosing).

A limitation of this study is that includes a small, heterogeneous sample. While this study aims to capture the real-life efficacy of T+C, the true effect of T/C is difficult to capture due to confounding factors from varying vaccination timeline, SARS-CoV-2 infection status, and other monoclonal antibody injections. Another limitation is the lack of long-term follow-up to assess rates of clinical breakthrough and safety. The study relied on participant surveys sent out a week after T+C doses and unsolicited patient self-reported to assess the safety and clinical breakthrough. Considering that preliminary safety and breakthrough data revealed events months after injections, future studies should be conducted on the long-term safety and breakthrough rate of T+C, particularly in SOTRs as their interaction with T+C has been understudied.

T+C is a reasonable complementary strategy to vaccination in high-risk SOTRs to improve neutralizing capacity against select Omicron sublineages. Assessment of the real-world efficacy and safety of T+C in SOTR is of utmost importance, since SOTRs face heightened risks of severe illness or death from COVID-19 and since only few SOTRs were included in the PROVENT trial. Studies such as ours incorporating both clinical and laboratory data offer valuable insights into the real-world efficacy and safety of T+C in one of our most vulnerable population. Further research examining the durability of neutralization against emerging Omicron sublineages is necessary, especially with evolving mutations and potential immune evasion ^9,16^.

## Data Availability

All data produced in the present study are available upon reasonable request to the authors

## Abbreviations

150+150mg T+C: 150mg Tixagevimab and 150mg Cilgavimab
300+300mg T+C: 300mg Tixagevimab and 300mg Cilgavimab
Anti-N: anti-nucleocapsid
AU: arbitrary units
BAU: binding antibody units
EIA: Enzyme Immunoassay
FDA: US Food and Drug Administration
IQR: interquartile range
MMF: mycophenolate
mRNA: messenger RNA
MSD: Meso Scale Diagnostics (MSD)
PrEP: pre-exposure prophylaxis
RBD: Receptor Binding Domain
SARS-CoV-2: severe acute respiratory syndrome coronavirus 2
SOTR: solid organ transplant recipients
T+C: Tixagevimab and Cilgavimab
ULOQ: upper limit of quantification
VOC: variants of concern (VOC)

## ACKNOWLEDGMENTS

The authors thank the participants of the Johns Hopkins COVID-19 Transplant Vaccine Study, without whom this research could not be possible. They also thank the members of the study team, including Brian J. Boyarsky MD, PhD; Laura Zeiser, MS; Chunyi Xia, BS; Kim Hall; Mary Sears, BS; Alex Alex; Jonathan Susilo.

## FUNDING/GRANT/AWARD INFORMATION

This work was supported by the Ben-Dov family, the Trokhan Patterson family, The ASTS Fryer Resident Scientist Award (Dr. Mitchell), grants 5T32DK007713 (Dr. Alejo), T32DK007732 (Dr. Chang), and K01DK101677 (Dr. Massie) from the National Institute of Diabetes and Digestive and Kidney Diseases; grant K24AI144954 (Dr. Segev), K08AI156021 (Dr. Karaba), U01AI138897, 3U01AI138897-04S1, and K23AI157893 (Dr. Werbel) from the National Institute of Allergy and Infectious Diseases; grant U54CA260491 (Dr. Pekosz, Dr. Karaba) from the National Cancer Institute.

---

**Table S1.**
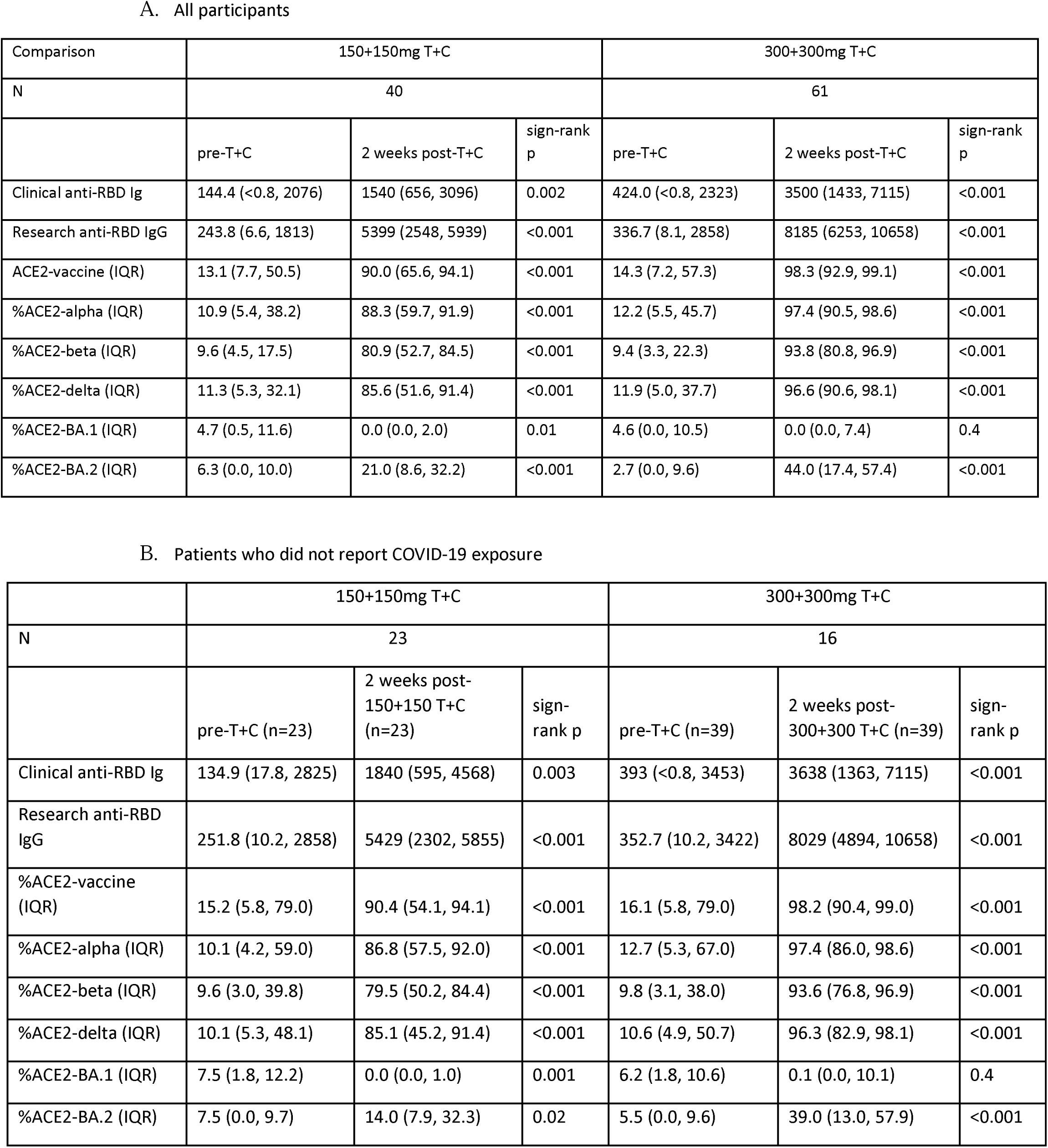
Median Anti-RBD titers and %ACE2 inhibition of SARS-CoV-2 variants, by Tixagevimab and Cilgavemab dose

A. All participants
| Comparison | 150+150mg T+C |  |  | 300+300mg T+C |  |  |
| --- | --- | --- | --- | --- | --- | --- |
| N | 40 |  |  | 61 |  |  |
|  | pre-T+C | 2 weeks post-T+C | sign-rank p | pre-T+C | 2 weeks post-T+C | sign-rank p |
| Clinical anti-RBD Ig | 144.4 (<0.8, 2076) | 1540 (656, 3096) | 0.002 | 424.0 (<0.8, 2323) | 3500 (1433, 7115) | <0.001 |
| Research anti-RBD IgG | 243.8 (6.6, 1813) | 5399 (2548, 5939) | <0.001 | 336.7 (8.1, 2858) | 8185 (6253, 10658) | <0.001 |
| ACE2-vaccine (IQR) | 13.1 (7.7, 50.5) | 90.0 (65.6, 94.1) | <0.001 | 14.3 (7.2, 57.3) | 98.3 (92.9, 99.1) | <0.001 |
| %ACE2-alpha (IQR) | 10.9 (5.4, 38.2) | 88.3 (59.7, 91.9) | <0.001 | 12.2 (5.5, 45.7) | 97.4 (90.5, 98.6) | <0.001 |
| %ACE2-beta (IQR) | 9.6 (4.5, 17.5) | 80.9 (52.7, 84.5) | <0.001 | 9.4 (3.3, 22.3) | 93.8 (80.8, 96.9) | <0.001 |
| %ACE2-delta (IQR) | 11.3 (5.3, 32.1) | 85.6 (51.6, 91.4) | <0.001 | 11.9 (5.0, 37.7) | 96.6 (90.6, 98.1) | <0.001 |
| %ACE2-BA.1 (IQR) | 4.7 (0.5, 11.6) | 0.0 (0.0, 2.0) | 0.01 | 4.6 (0.0, 10.5) | 0.0 (0.0, 7.4) | 0.4 |
| %ACE2-BA.2 (IQR) | 6.3 (0.0, 10.0) | 21.0 (8.6, 32.2) | <0.001 | 2.7 (0.0, 9.6) | 44.0 (17.4, 57.4) | <0.001 |

B. Patients who did not report COVID-19 exposure
|  | 150+150mg T+C |  |  | 300+300mg T+C |  |  |
| --- | --- | --- | --- | --- | --- | --- |
| N | 23 |  |  | 16 |  |  |
|  | pre-T+C (n=23) | 2 weeks post-150+150 T+C (n=23) | sign-rank p | pre-T+C (n=39) | 2 weeks post-300+300 T+C (n=39) | sign-rank p |
| Clinical anti-RBD Ig | 134.9 (17.8, 2825) | 1840 (595, 4568) | 0.003 | 393 (<0.8, 3453) | 3638 (1363, 7115) | <0.001 |
| Research anti-RBD IgG | 251.8 (10.2, 2858) | 5429 (2302, 5855) | <0.001 | 352.7 (10.2, 3422) | 8029 (4894, 10658) | <0.001 |
| %ACE2-vaccine (IQR) | 15.2 (5.8, 79.0) | 90.4 (54.1, 94.1) | <0.001 | 16.1 (5.8, 79.0) | 98.2 (90.4, 99.0) | <0.001 |
| %ACE2-alpha (IQR) | 10.1 (4.2, 59.0) | 86.8 (57.5, 92.0) | <0.001 | 12.7 (5.3, 67.0) | 97.4 (86.0, 98.6) | <0.001 |
| %ACE2-beta (IQR) | 9.6 (3.0, 39.8) | 79.5 (50.2, 84.4) | <0.001 | 9.8 (3.1, 38.0) | 93.6 (76.8, 96.9) | <0.001 |
| %ACE2-delta (IQR) | 10.1 (5.3, 48.1) | 85.1 (45.2, 91.4) | <0.001 | 10.6 (4.9, 50.7) | 96.3 (82.9, 98.1) | <0.001 |
| %ACE2-BA.1 (IQR) | 7.5 (1.8, 12.2) | 0.0 (0.0, 1.0) | 0.001 | 6.2 (1.8, 10.6) | 0.1 (0.0, 10.1) | 0.4 |
| %ACE2-BA.2 (IQR) | 7.5 (0.0, 9.7) | 14.0 (7.9, 32.3) | 0.02 | 5.5 (0.0, 9.6) | 39.0 (13.0, 57.9) | <0.001 |

**Table S2.** Proportion of participants achieving ≥20% ACE2 inhibition before and two weeks after 300-mg of Tixagevimab plus Cilgavimab, by SARS-CoV-2 variant

A. All participants
| Variants of concern | Pre-T+C | 2 weeks post-T+C | Exact McNemar (p-value) |
| --- | --- | --- | --- |
| Vaccine (ancestral, WA/1) | 28/61 (46%) | 61/61 (100%) | <0.001 |
| Alpha (B.1.1.7) | 27/61 (44%) | 61/61 (100%) | <0.001 |
| Beta (B.1.351) | 16/61 (26%) | 61/61 (100%) | <0.001 |
| Delta (B.1.617.2) | 24/61 (39%) | 61/61 (100%) | <0.001 |
| Omicron BA.1 | 5/61 (8%) | 10/61 (16%) | 0.06 |
| Omicron BA.2 | 4/61 (7%) | 44/61 (72%) | <0.001 |

B. Participants without reported recent SARS-CoV-2 antigen exposure
| Variants of concern | Pre-T+C | 2 weeks post-T+C | Exact McNemar (p-value) |
| --- | --- | --- | --- |
| Vaccine (ancestral, WA/1) | 17/39 (44%) | 39/39 (100%) | <0.001 |
| Alpha (B.1.1.7) | 17/39 (44%) | 39/39 (100%) | <0.001 |
| Beta (B.1.351) | 12/39 (31%) | 39/39 (100%) | <0.001 |
| Delta (B.1.617.2) | 15/39 (38%) | 39/39 (100%) | <0.001 |
| Omicron BA.1 | 3/39 (8%) | 6/39 (15%) | 0.25 |
| Omicron BA.2 | 2/39 (5%) | 26/39 (67%) | <0.001 |

**Table S3.** Median %ACE2 inhibition of participants achieving ≥20% ACE2 inhibition before and two weeks after 300-mg of Tixagevimab plus Cilgavimab, by SARS-CoV-2 variant

A. All participants
| Variants of concern | Pre-T+C (%) | 2 weeks post-T+C (%) |
| --- | --- | --- |
| Vaccine (ancestral, WA/1) | 69.3 (43.6, 91.8) (n=28) | 98.3 (92.9, 99.1) (n=61) |
| Alpha (B.1.1.7) | 59.0 (33.5, 83.6) (n=27) | 97.4 (26.5, 67.2) (n=61) |
| Beta (B.1.351) | 47.8 (36.1, 68.4) (n=16) | 93.8 (80.8, 96.9) (n=61) |
| Delta (B.1.617.2) | 53.0 (32.1, 78.0) (n=24) | 96.6 (90.6, 98.1) (n=61) |
| Omicron BA.1 | 29.6 (26.5, 67.2) (n=5) | 37.0 (23.6, 56.9) (n=10) |
| Omicron BA.2 | 41.4 (29.8, 56.8) (n=4) | 51.7 (39.7, 63.0) (n=44) |

B. Participants without reported recent SARS-CoV-2 antigen exposure
| Variants of concern | Pre-T+C (%) | 2 weeks post-T+C (%) |
| --- | --- | --- |
| Vaccine (ancestral, WA/1) | 79.4 (51.5, 91.1) (n=17) | 98.2 (90.4, 99.0) (n=39) |
| Alpha (B.1.1.7) | 68.3 (40.6, 80.3) (n=17) | 97.4 (86.0, 98.6) (n=39) |
| Beta (B.1.351) | 47.8 (38.9, 63.1) (n=12) | 93.6 (76.8, 96.9) (n=39) |
| Delta (B.1.617.2) | 59.9 (41.4, 76.6) (n=15) | 96.3 (82.9, 98.1) (n=39) |
| Omicron BA.1 | 26.5 (21.3, 29.6) (n=3) | 29.5 (23.6, 40.7) (n=6) |
| Omicron BA.2 | 29.8 (26.3, 33.3) (n=2) | 52.6 (39.0, 60.5) (n=26) |

**Table S4.**
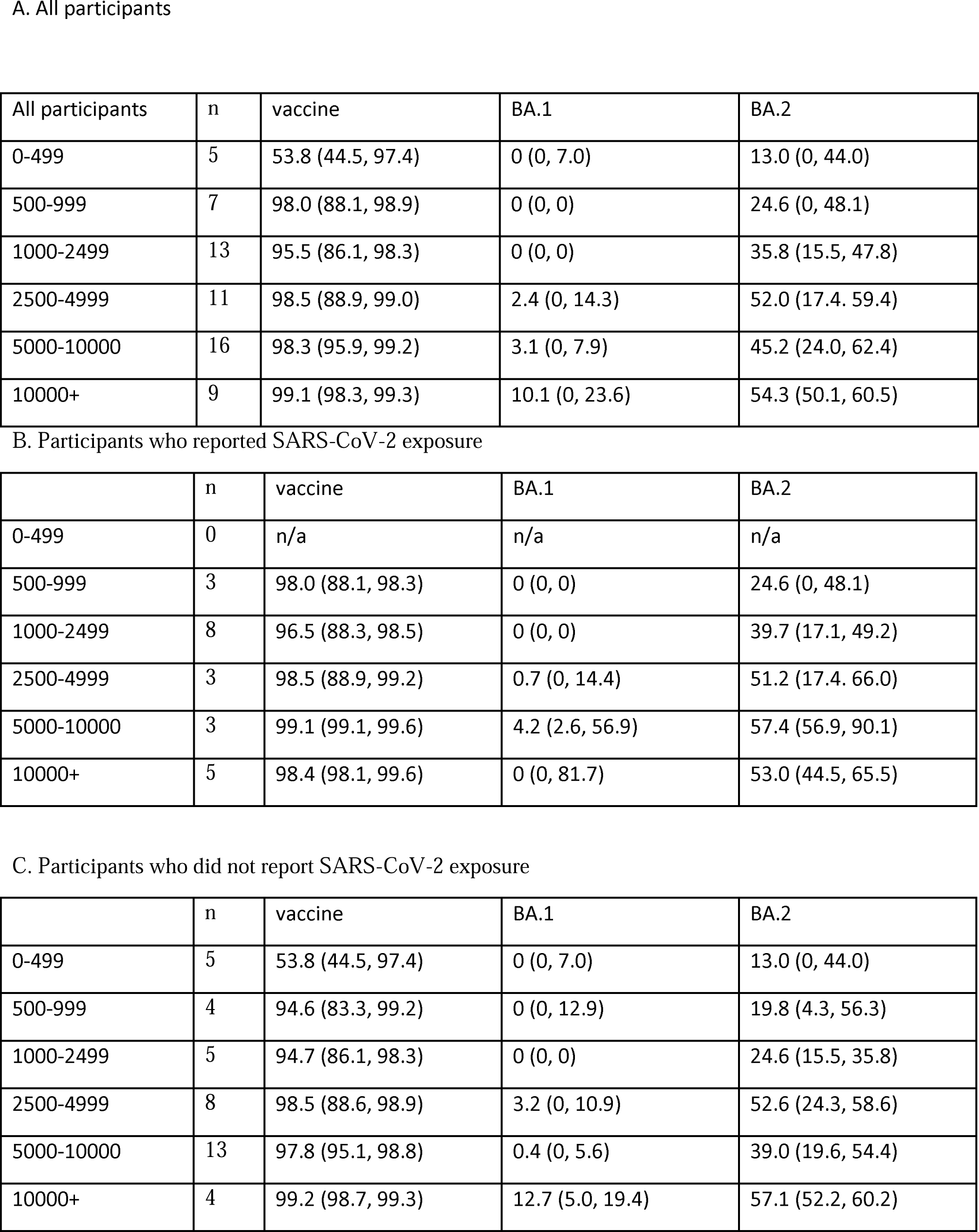
Proportion of participants achieving neutralizing inhibition at select anti-RBD thresholds after 300-mg of Tixagevimab plus Cilgavimab, by SARS-CoV-2 variant

**Figure S1.**
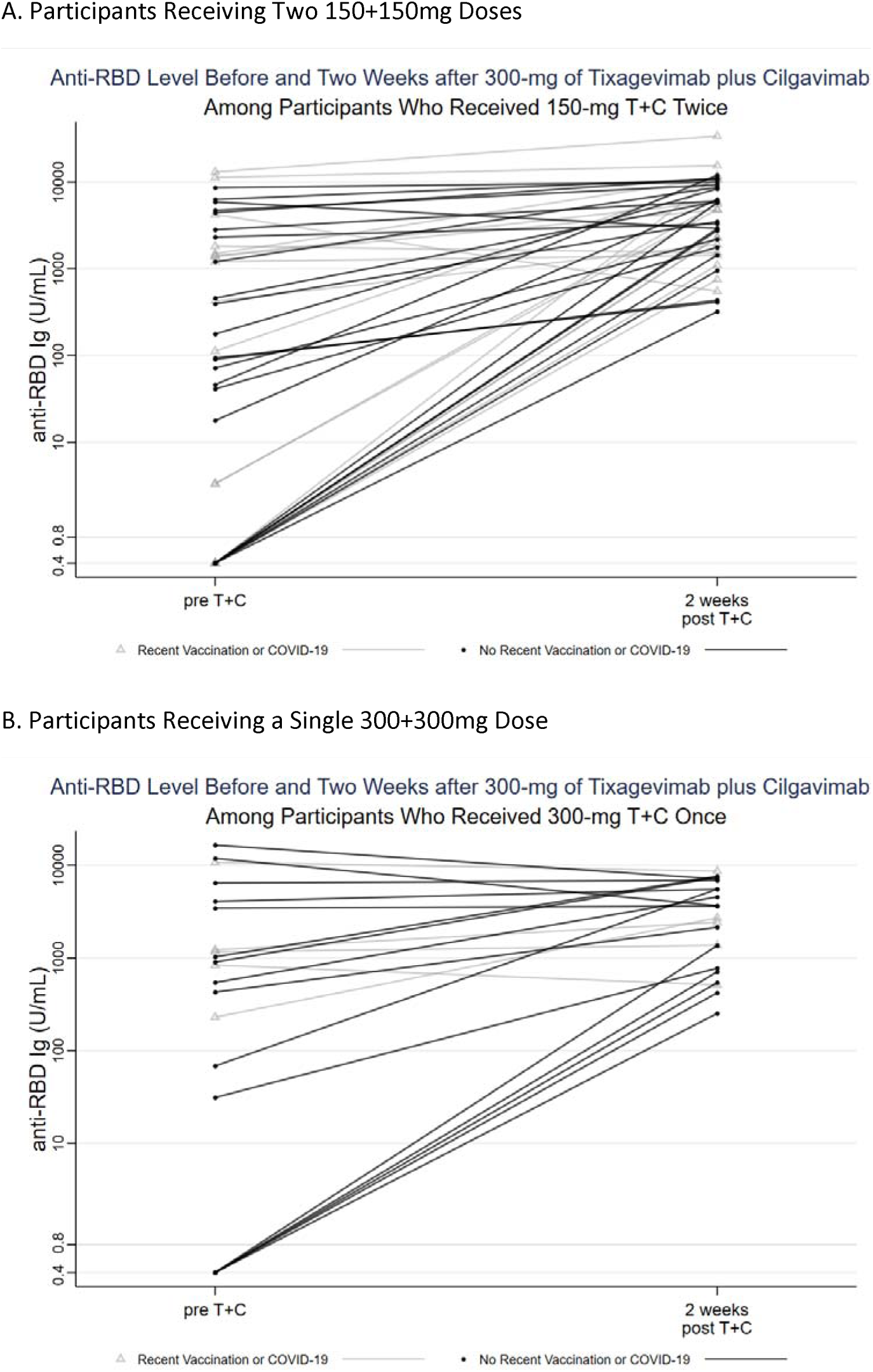
Anti-RBD level before and two weeks after 300+300mg of Tixagevimab plus Cilgavimab, by dose administration

**Figure S2.**
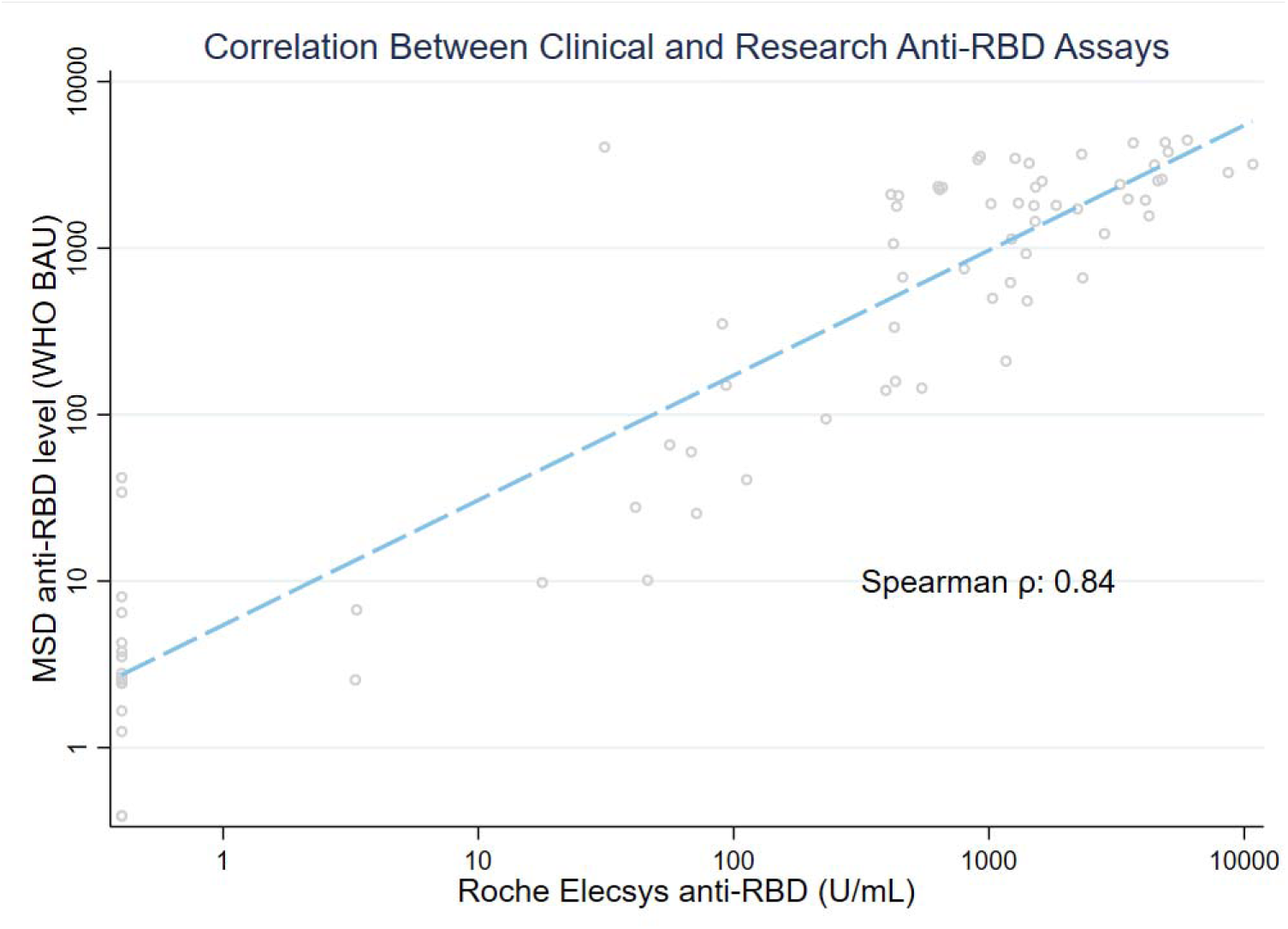
Correlation between clinical and research anti-RBD assays in vaccinated SOTRs following Tixagevimab and Cilgavimab injection Correlation was performed only for samples between upper and lower limit of detection on both assays (76 samples, n=51); if using all samples: ρ = 0.65 (176 samples, n=61); if limited to post-injection samples below the upper limited of quantification: ρ = 0.65 (26 samples, n=15); if using all post-injection samples: ρ = 0.43 (115 samples, n=61)

**Figure S3.**
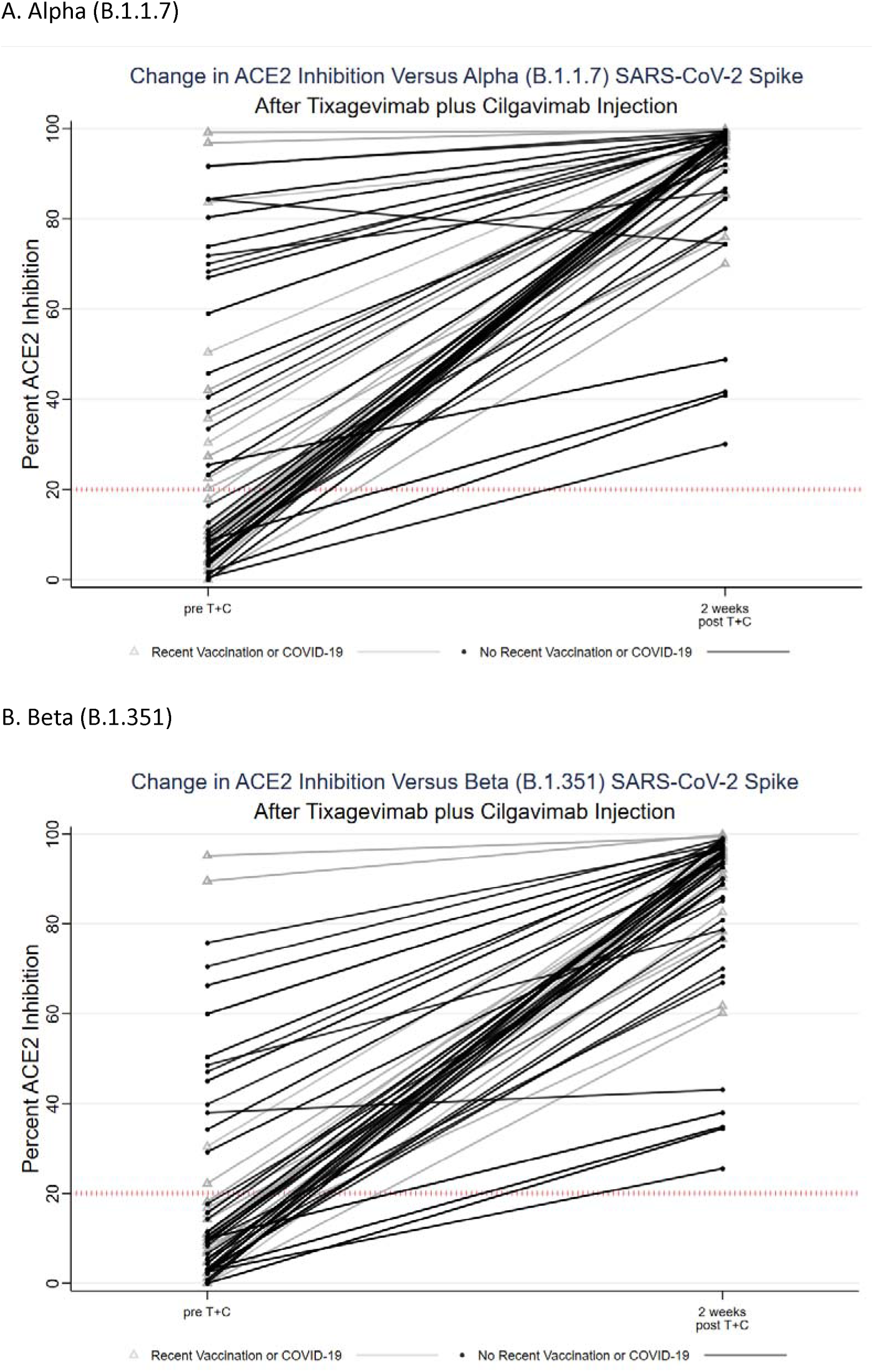
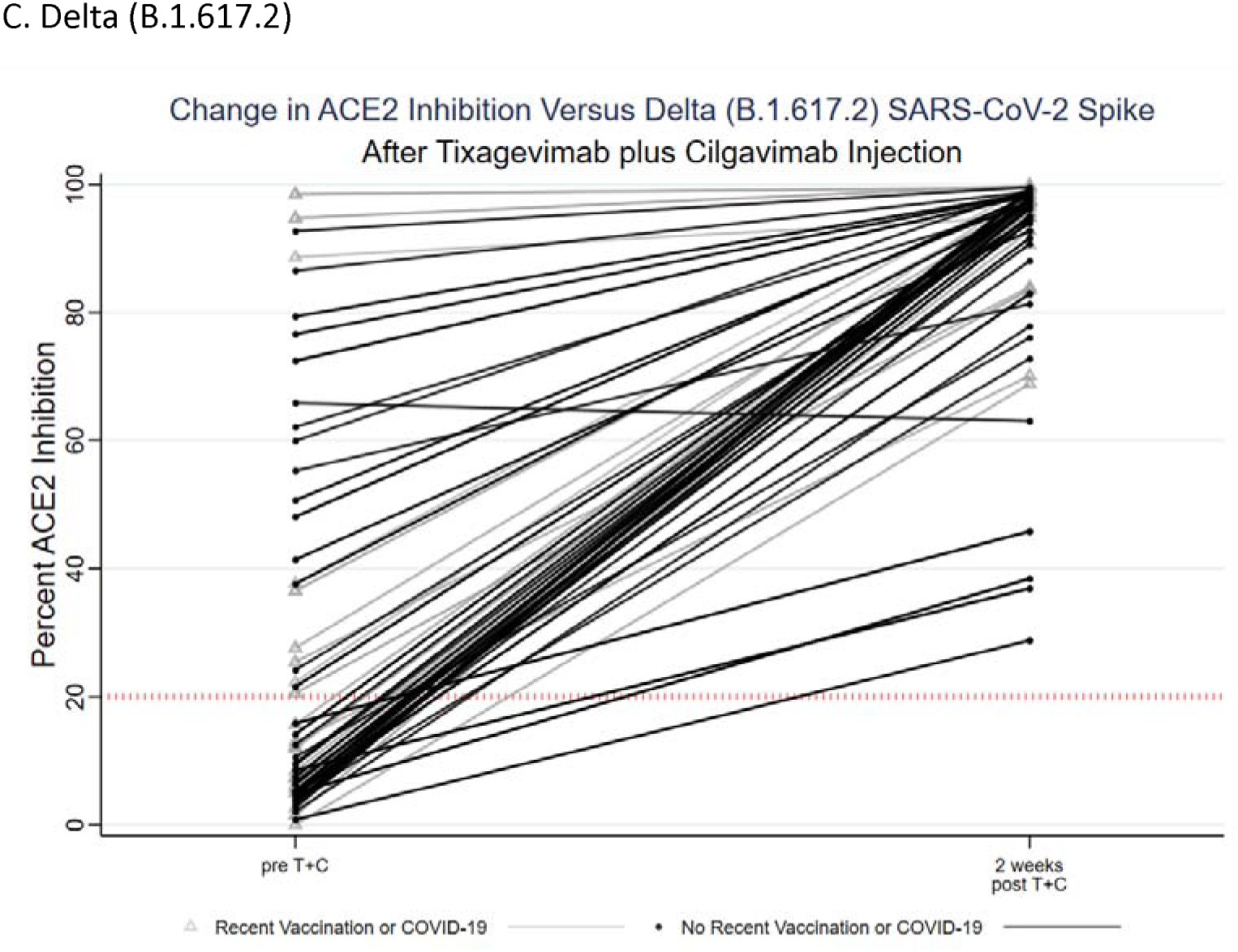
Change in ACE2 inhibition versus SARS-CoV-2 variants after Tixagevimab plus Cilgavimab Injection

**Figure S4.**
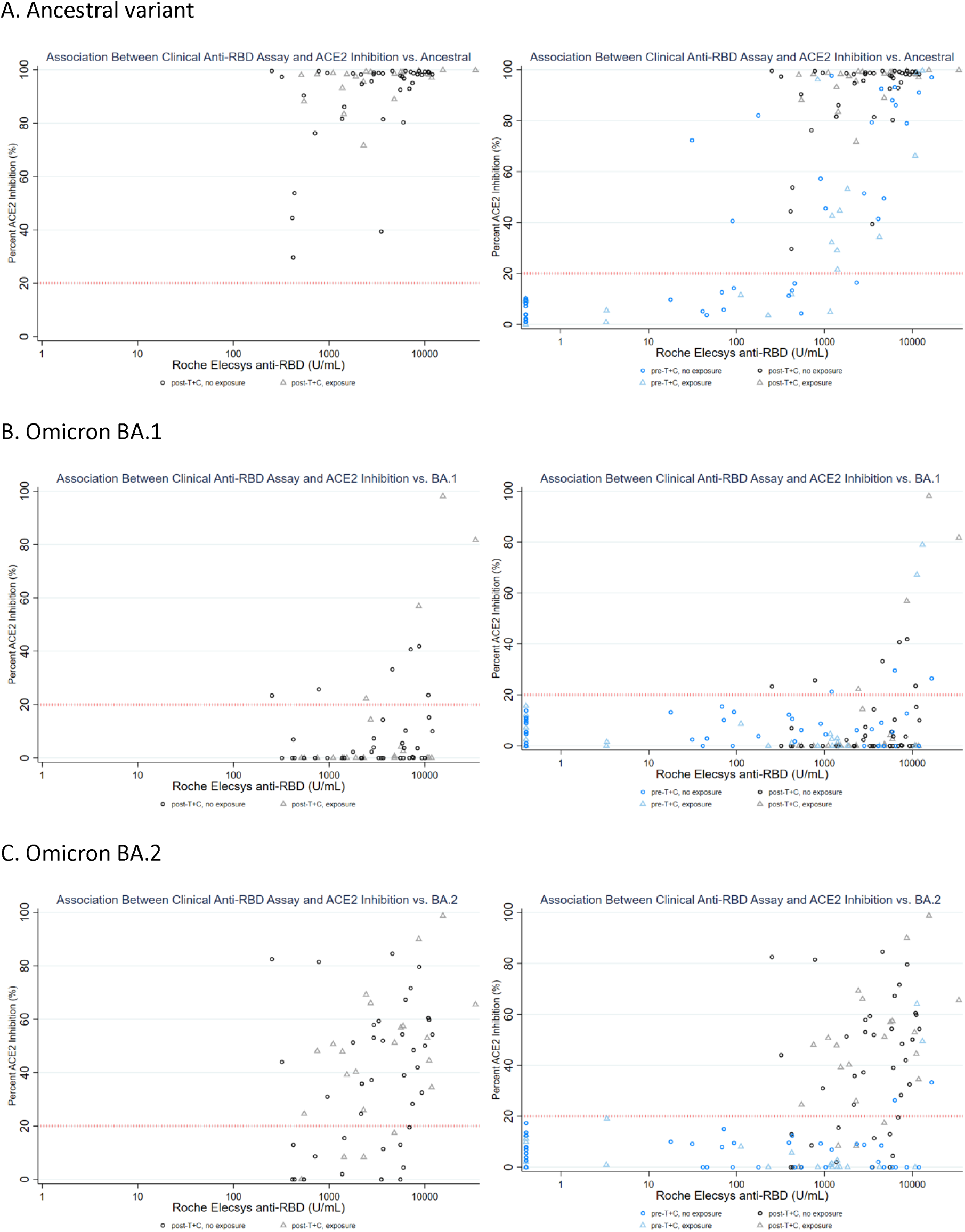
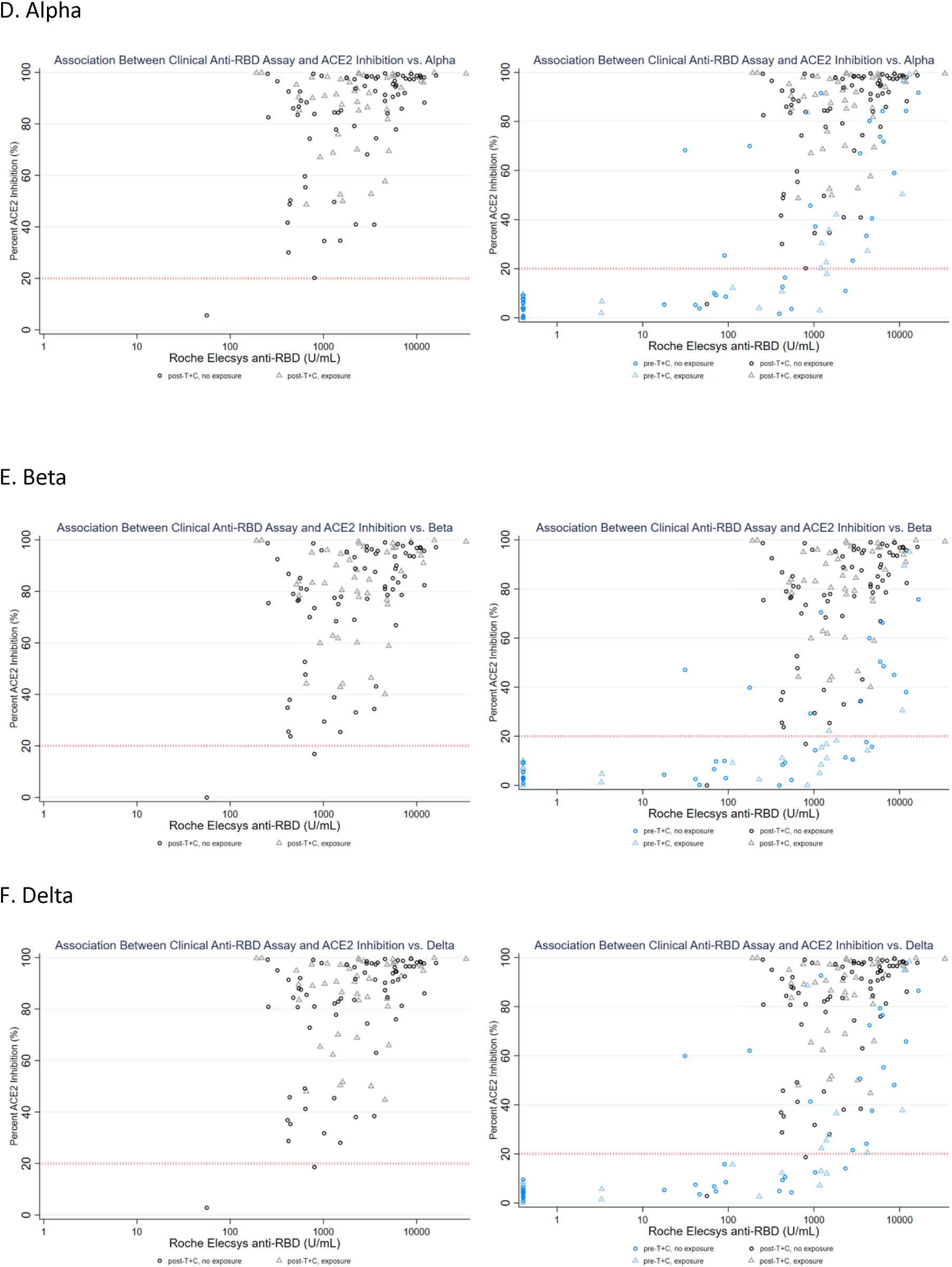
Association between surrogate neutralization of SARS-CoV-2 variants and anti-RBD level before and after Tixagevimab and Cilgavimab Injection Increasing anti-RBD titer after T+C moderately correlated with increasing %ACE2 inhibition (ρ=0.43) against the vaccine strain. Using all 176 samples from 61 participants before and after injection, the correlation between anti-RBD and %ACE2 inhibition against BA.1: ρ=0.04, BA.2: ρ=0.40, alpha: ρ=0.62, beta: ρ=0.61, delta: ρ=0.62. Using only the 115 post-T+C samples from 61 participants, the correlation between anti-RBD and %ACE2 inhibition against BA.1: ρ=0.32, BA.2: ρ=0.39, alpha: ρ=0.40, beta: ρ=0.41, delta: ρ=0.41

**Figure S5.**
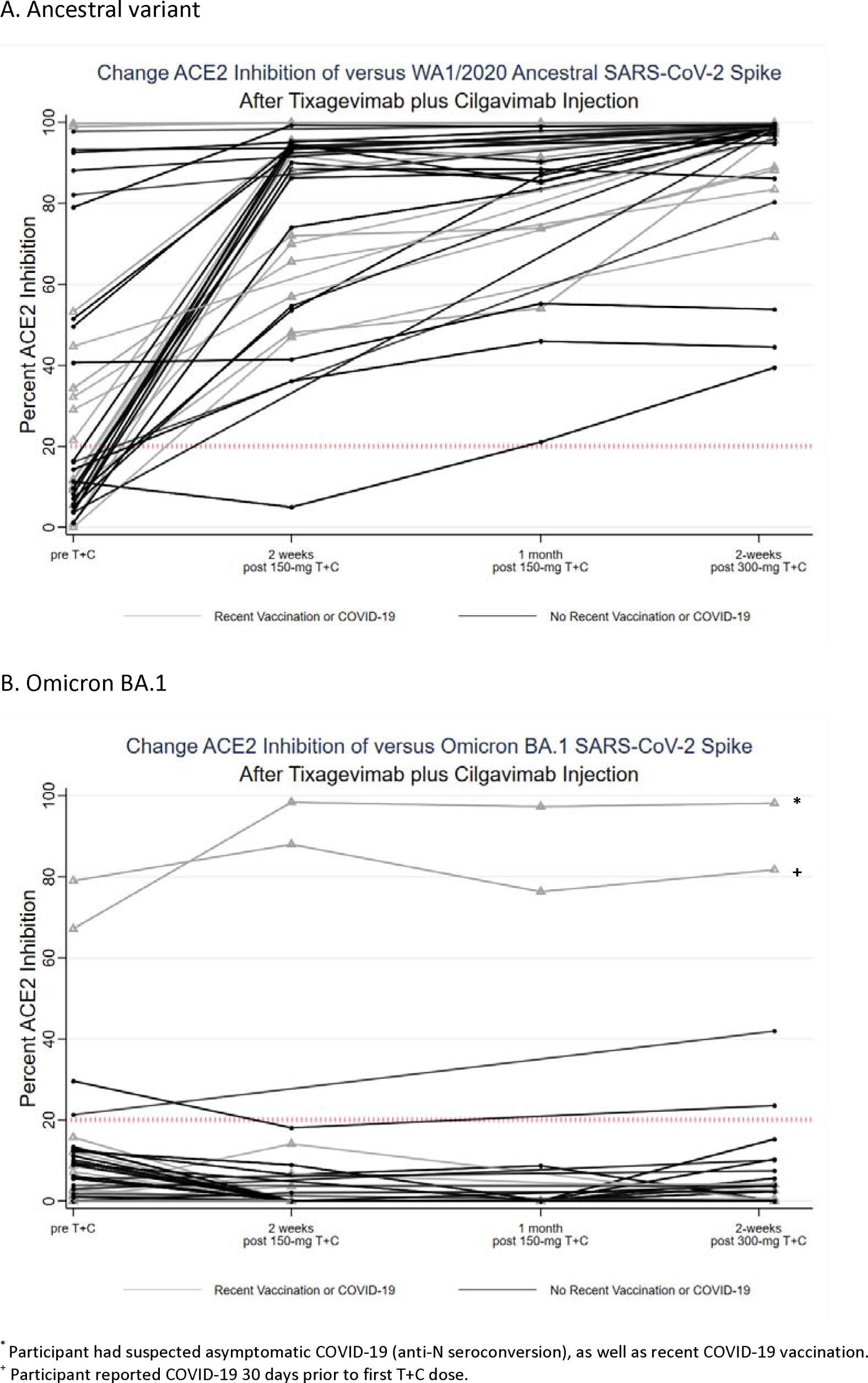
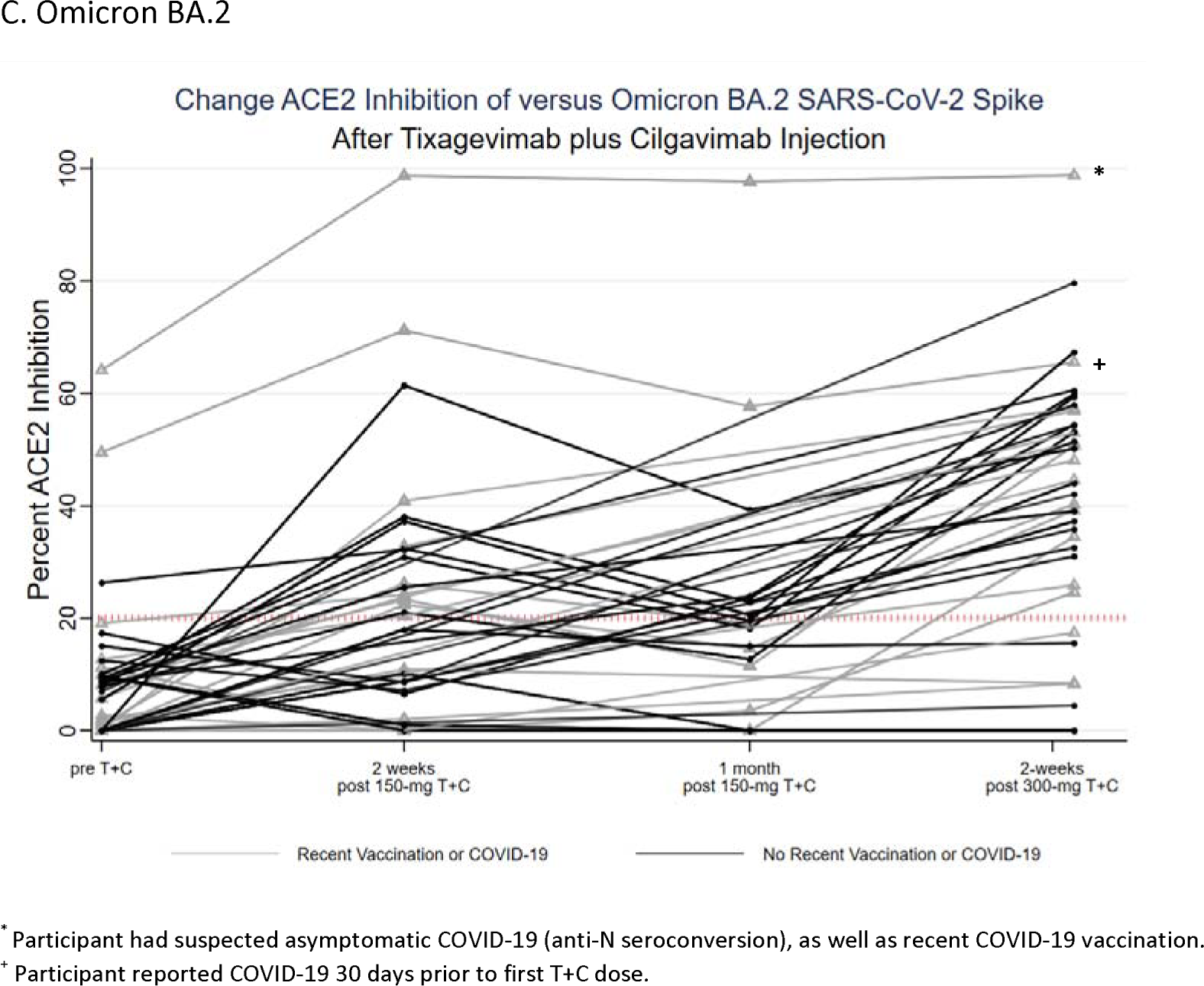
Change in ACE2 inhibition versus SARS-CoV-2 variants following first and second 150+150mg doses of Tixagevimab plus Cilgavimab

**Figure S6.**
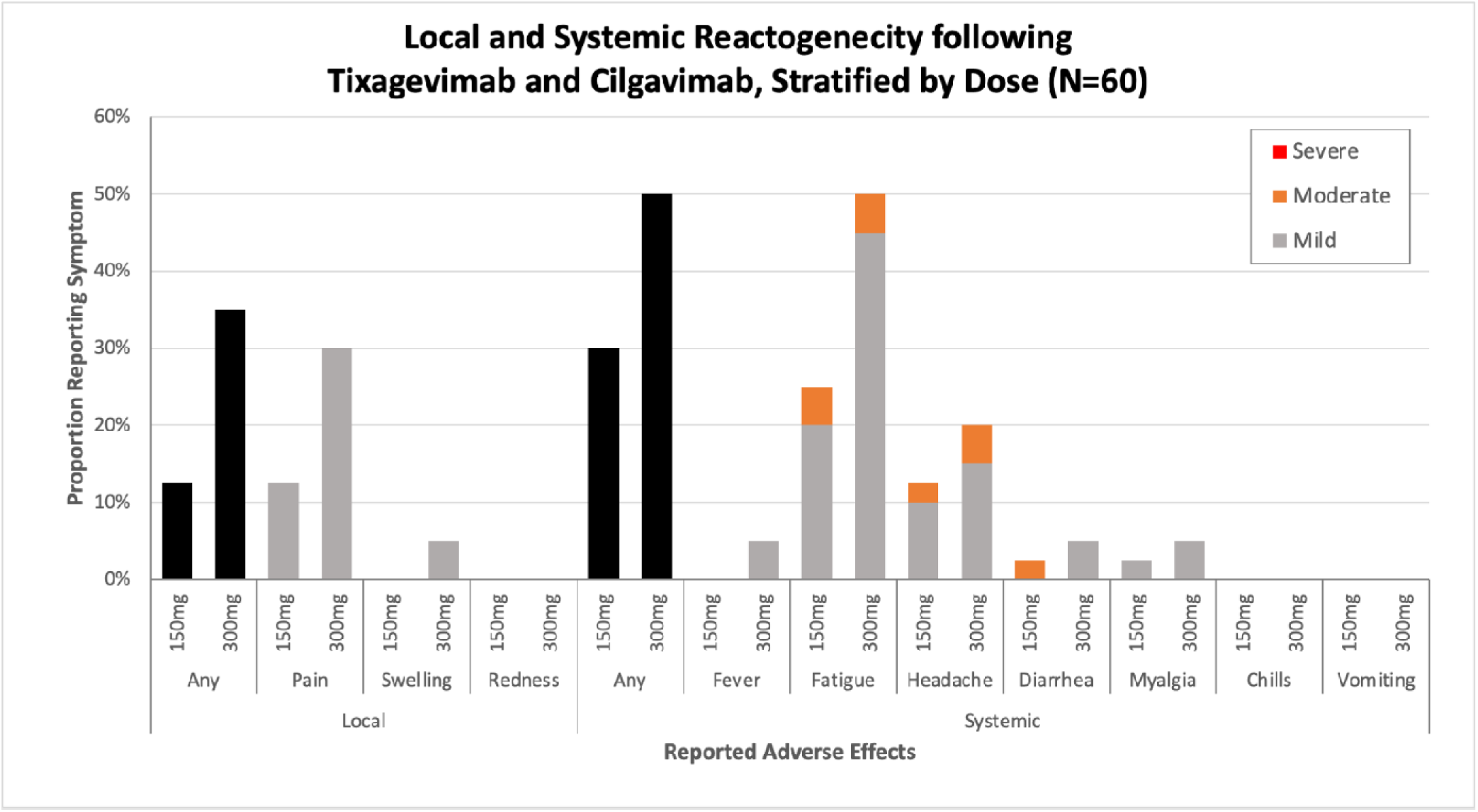
Self-reported local and systemic adverse reactions after Tixagevimab plus Cilgavimab Injection, by dose administration. Responses were received from 60/61 (98%) of participants. Some surveys were incomplete in documenting degree of redness (n=1), fatigue (n=1), myalgia (n=2).

